# MDAtlas: A Multiomics Meniere Disease Database of Genes and Cells

**DOI:** 10.1101/2025.10.03.25336856

**Authors:** Kiana Bagheri-Lotfabad, Pablo Cruz-Granados, Muhammad Umair, Alberto M. Parra-Perez, Sayedali Mohseni, Mai T. Pham, Giselle Bianco-Bortoletto, Sung Huhn Kim, Rick A. Friedman, Daniel Burn, Jose A. Lopez-Escamez, the Meniere Disease Multiomic Aggregation Consortium (MD-MAC)

## Abstract

The MDAtlas is a crowdsourcing initiative to provide an open access resource with multiomic dataset about Meniere Disease (MD) -a debilitating audiovestibular disorder- to the scientific/medical community. The platform (https://multiomic-md.sydney.edu.au) integrates anonymised genomic, transcriptomic and epigenomic datasets from several MD cohorts with diverse ancestry through the MD multiomic Agreggation Consortium (MD-MAC), helping users investigate common and rare genomic variability, immune cell gene expression, and DNA methylation patterns involved in these conditions. MDAtlas supports secure data upload, advanced filtering, and easy downloading, allowing researchers to compare datasets across populations and discover new disease mechanisms or biomarkers for diagnosis and therapy. By providing interactive tools and harmonized data, MDAtlas accelerates scientific progress and collaboration in hearing and vestibular research.

## Introduction

Meniere Disease (MD) is a debilitating inner ear disorder characterised by episodic vertigo, fluctuating sensorineural hearing loss, tinnitus, and aural fullness[1,2] The condition is associated with an accumulation of endolymph in the cochlear duct, termed endolymphatic hydrops, though the molecular mechanisms linking hydrops to symptom occurrence remain unclear [2,3]. Five clinical phenotypes have been identified based on clinical predictors in both unilateral (UMD) and bilateral (BMD) MD, that exhibit significant phenotypic heterogeneity, that may involve comorbidities such as migraine and autoimmune disorders [4,5]. Furthermore, most MD patients show systemic inflammation creating a challenging diagnosis and management for clinicians [6].

Familial MD (FMD) is found in 10% of cases and includes monogenic and polygenic (biallelic) inheritance with autosomal dominant or recessive patterns [3]. Pathogenic variants have been identified in more than 20 genes, including *OTOG, TECTA* and *GJD3* in European FMD cases [3,7]. These genes encode structural proteins of the tectorial and otolithic membranes in the inner ear sensory epithelia, which are essential for hearing and balance [3,8,9]. Around 15% FMD cases involve two heterozygous variants in *OTOG*, which can disrupt the stability of the tectorial membrane and affect how signals are transmitted in the ear. Rare missense and Loss-of-function (LoF) variants in *TECTA* have also been found in several MD families, suggesting that mutations in this gene may contribute to the disease pathophysiology [10]. Moreover, several familial and non-familial MD patients share a complex haplotype in the *GJD3* gene; this gene encodes a gap junction protein that helps with cell-to-cell communication in the inner ear, and missense variant in this haplotype may interfere with the connexon assembly and contribute to MD [7].

Studies employing cytokine profiling, bulk RNA sequencing (RNAseq) and single-cell RNA sequencing (scRNAseq) have identified several immunophenotypes in sporadic MD (SMD) based on cytokine levels and systemic inflammation [11–14]. One immune phenotype is characterised by high levels of IL-1β and linked to autoinflammation [11,14]. A second phenotype is driven by overexpression of IgE and Th2 cytokines, with an allergy-like immune response [13,15]. The third phenotype is autoimmune MD (AIMD) characterised by the presence of autoantibodies [4,5]. Notably, a longitudinal study that followed individuals over a two-year period demonstrated that cytokine profiles remain stable over time within each immunophenotype [6]. In addition, recent advances have integrated transcriptomic and genomic data to identify rare variants in immune-related genes. A study reported that patients with the autoinflammatory phenotype carry a variant in the *KIF1B* gene, which may contribute to immune system dysfunction [16].

With the increasingly use of Next-Generation Sequencing (NGS) data in the study of hearing and vestibular disorders, several databases have been developed to consolidate relevant information. Resources such as Deafness Variation Database (DVD) [17], gEAR Portal [18], Gene4HL [19] and DVPred [20] provide curated data specific to audiovestibular disorders. Additionally, population- or tissue-level multiomics data are available through databases like gnomAD [21], Spanish Collaborative Variant Server (CSVS) [22], GTEx Project [23]. Despite these advances, MD human molecular data remain fragmented by isolated genomic, epigenomic, and transcriptomic datasets. This paper introduces the *MDAtlas: Genes and Cells*, a multiomics platform integrating aggregated variant data, Cytosine-phosphate-Guanine (CpG) methylation profiles, and RNA-seq and scRNAseq from different MD cohorts (https://multiomic-md.sydney.edu.au/). This open resource will serve as reference dataset to facilitate multiomic analysis, uncover novel MD-specific mechanisms, resolve phenotypic heterogeneity, and enable personalised diagnostics. By harmonising dispersed datasets and enabling user-driven comparisons, this resource addresses critical gaps identified in familial and sporadic MD genetics, paving the way for mechanistic insights and therapeutic discovery.

### Database Implementation

To deliver a powerful and scalable platform, the MDAtlas architecture is built as a comprehensive, full-stack web application designed for performance, reliability and maintainability. The entire system is containerised using Docker, which ensures a consistent environment from development to production. This architecture consists of three core components: a React.js frontend, a Node.js backend, and a MongoDB database [24,25].

The front-end user interface is developed using React.js, creating a responsive and dynamic experience for data exploration and visualisation. The user interface is a dynamic and responsive single-page application (SPA) developed with React.js. Its component-based structure promotes reusability and clean separation of concerns. This is evident in our codebase, which includes specialised components for data visualisation (e.g., GenomicTable.jsx, SingleCellTranscriptomicTable.jsx) and user experience elements like the ProgressOverlay for loading states. Global application state is managed efficiently using Redux, which provides a predictable state container for a seamless user journey through complex data filtering and analysis tasks [25].

Our back-end services are built on Node.js with the Express.js framework, providing a robust RESTful API to handle data management, user authentication, and analysis requests. The backend follows a standard structure with dedicated directories for routes (API endpoints), controllers (business logic), and models (database schemas), making the system logical and easy to maintain.

For our database, we have implemented MongoDB. Its NoSQL, document-based structure offers the flexibility required to store the complex and diverse metadata associated with multiomic studies [24]. The entire platform is deployed on Amazon Web Services (AWS) and leverages The University of Sydney’s CI/CD pipelines. This automated process ensures that any code committed to the develop, release/test, or main branches is automatically built, tested, and deployed, ensuring stable and consistent updates [26]. Users can sign up and access data using either a standard email registration or their ORCID account (https://orcid.org/) for seamless integration.

### Database content and usage

The MDAtlas database is a highly specialised digital library, meticulously curated to centralise critical molecular data for MD and hearing loss research. It houses a collection of genomic, epigenomic, and transcriptomic data from 467 MD participants, generated by collaborators in Spain, Australia, the US, Brazil and South Korea covering Non-Finish European (NFE), East-Asian (EAS) and Admixed American (AMR) populations (Figure 1).

**Figure 1.**
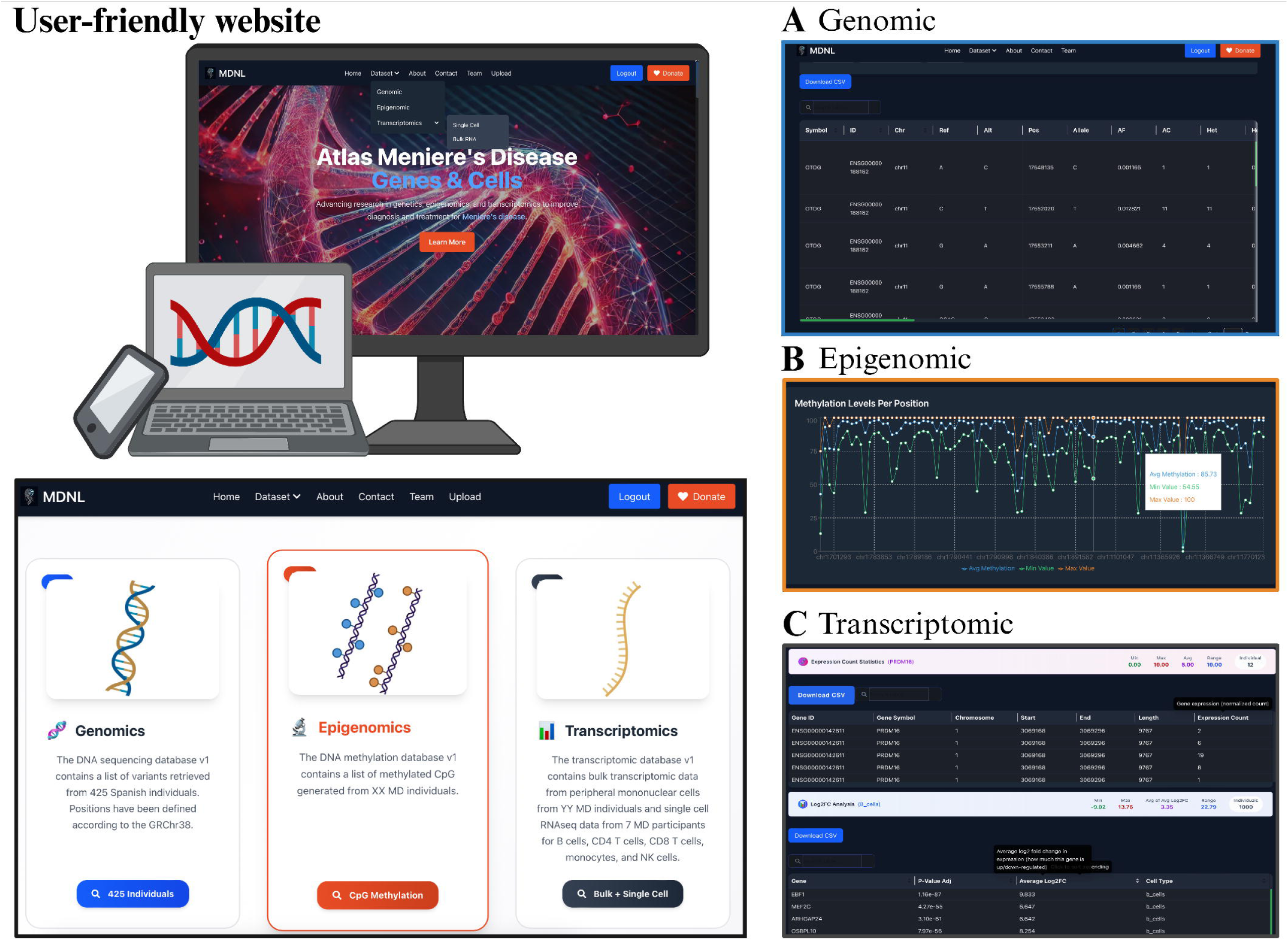
MDAtlas architecture and data model. The MDAtlas website provides an interactive platform to explore Meniere disease multi-omics datasets to Meniere’s disease. The genomic dataset allows researchers to select populations (Brazil, Korea, Spain), apply filters, and view detailed variant information (e.g., gene symbol, chromosome location, allele frequencies, functional impact), with column-specific descriptions available on hover. Filtered results can be downloaded for further analysis. The epigenomic dataset provides DNA methylation profiles, which can be filtered and downloaded, with interactive visualisation of methylation levels per genomic position based on user-defined parameters. The transcriptomic dataset includes both bulk and single-cell RNA sequencing data, enabling users to filter by conditions, view summary statistics such as minimum, maximum, and average expression values, and download customised subsets of gene expression data.

**Figure 2.**
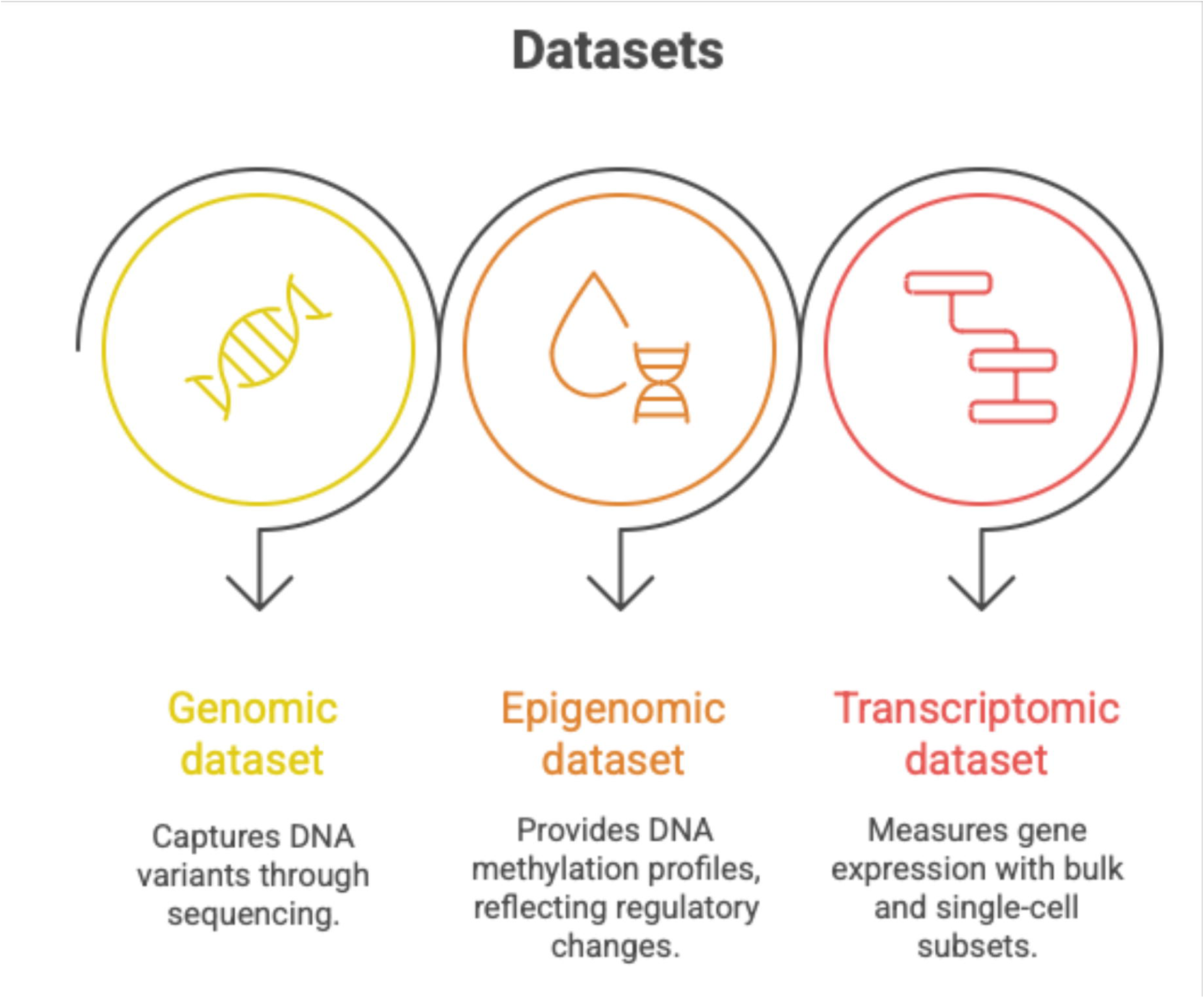
Multiomics datasets available on the website. The MDAtlas includes three datasets: genomics, epigenomics and transcriptomics data. The genomic dataset captures DNA sequence variants, enabling the identification of genetic alterations. The epigenomic dataset provides DNA methylation profiles, reflecting regulatory changes in gene activity. The transcriptomic dataset quantifies gene expression using both bulk and single-cell RNA sequencing, offering insights into cellular heterogeneity.

To make this complex information accessible, the platform provides dedicated ‘reading rooms’ or data browsers for each *omic* layer, allowing users to dive deep into specific biological data:

- **Genomic:** This dataset is composed of whole exome sequencing (WES) and whole genome sequencing (WGS) files from 467 individuals of NFE, AMR and EAS background (**Table 1A**). We have annotated our data against gnomAD v4.1.0 [21] to retrieve allelic frequency (AF) in per population. Users can compare the AF on their cohort, the MDAtlas cohorts and gnomAD to determine frequency of the variant.
- **Epigenomic:** This dataset is built on peripheral mononuclear cells (PBMC), and it allows users to look for differentially methylated CpG (DMC) sites in the Atlas cohort (**Table 1B)**. We have created a table that includes maximum, minimum and average percentage methylation for each DMC, that allows users to identify DMCs that may differentiate clusters of MD individuals.
- **Transcriptomic:** This dataset was built using RNAseq data from PBMCs and scRNAseq data covering B cells, CD4^+^ and CD8^+^ T cells, monocytes, and natural killer (NK) cells (**Table 1C**). This allows users to visualise transcript expression in specific cell types, as well as in the samples overall. As in the *Epigenomic* dataset, both RNAseq and scRNAseq datasets has a maximum, minimum and average counts or fold change, respectively, per transcript allowing to cluster MD cases. Furthermore, scRNAseq dataset has a *p*-value adjusted using the Bonferroni correction for statistical significance.

**Table 1.** Summary statistics of MDAtlas datasets. A) The genomic dataset includes three cohorts: 376 Non-Finnish European (NFE), 16 East Asian (EAS), and 22 Admixed American (AMR) individuals. It lists the total number of genes present, along with the counts of variants categorized by consequence (missense, synonymous, intronic, and loss-of-function [LoF], which includes frameshift variants, stop gained, splice donor and acceptor, stop lost, start lost and stop retained) and impact (high, moderate, low). **B)** The epigenomic dataset comprises 53 NFE individuals with data from peripheral blood mononuclear cells (PBMCs), including the number of genes and CpG sites. **C)** The transcriptomic datasets are generated from PBMCs of NFE individuals. The bulk transcriptomic dataset includes 12 individuals with raw counts, whereas the single-cell dataset comprises 7 individuals with pre-processed data (filtered using Seurat v5 for high mitochondrial gene cells and doublets, and annotated by cell type). The single-cell data consists of B cells, CD4+ T cells, CD8+ T cells, NK cells, and monocytes.

| Genomic Dataset |  |  |  |  |
| --- | --- | --- | --- | --- |
| Population |  | NFE | EAS | AMR |
| N |  | 376 | 16 | 22 |
| Genes |  | 31,766 | 37,722 | 71,730 |
| Variants | *Missense* | 115,666 | 46,328 | 20,352 |
|  | *Synonymous* | 84,839 | 45,153 | 21,466 |
|  | *Intronic* | 30,306 | 101,667 | 924,220 |
|  | *LoF* | 2,151 | 1,547 | 502 |
| Impact | *High* | 6,236 | 1,685 | 753 |
|  | *Moderate* | 122,414 | 48,733 | 21,054 |
|  | *Low* | 88,608 | 61,752 | 30,934 |

**B**
| Epigenomic Dataset |  |
| --- | --- |
| **Population** | NFE |
| **N** | 53 |
| **Tissue** | PBMC |
| **Genes** | 15,405 |
| **CpG** | 684,141 |

**C**
| Transcriptomic Dataset |  |  |  |
| --- | --- | --- | --- |
| **Population** | NFE |  |  |
| **Tissue** | PBMC |  |  |
| **Method** | Bulk | Single-Cell |  |
| **N** | 12 | 7 |  |
| **Preparation** | Raw | Pre-processed |  |
| **Genes** | 43,807 | *B Cells* | 16,405 |
|  |  | *CD4* | 15,134 |
|  |  | *CD8* | 15,002 |
|  |  | *NK Cells* | 15,985 |
|  |  | *Monocytes* | 14,180 |

Authenticated users can securely submit their own data, filter existing datasets using granular criteria, and download data packages for offline analysis. Users can use the platform for several key purposes:

- **Uploading Data:** Users can securely upload their own genomic datasets to the platform. This feature does not add their data to the public repository but rather allows them to use the Atlas tools to compare their cohort against our extensive datasets. It allows users to filter and prioritise variants for genetic diagnosis and research.
- **Filter and Explore:** The platform is equipped with a friendly interface that acts like a sophisticated search engine for data. Users can apply granular filters such as isolating variants within specific genomic coordinates, searching for a particular gene, pinpointing methylation sites, or exploring gene expression within PBMC subtypes.

This allows users to precisely narrow down the number of variants and genes to answer highly specific biological questions.

- **Download Data:** Once the data of interest has been identified, users can easily download it. The platform packages all the relevant data along with its complete metadata the crucial contextual information about the samples and experiments ensuring that the information is ready for deeper, offline analysis.

### MDAtlas Scheme

#### Platform Architecture and Data Scheme

MDAtlas is a full-stack web application with a clear separation of concerns, containerised using Docker for consistent development and deployment.

#### Application Architecture

The project follows a standard monorepo structure, organised as follows:

- app/frontend/: Contains the React.js single-page application. It uses Redux for global state management and features a component-based architecture with dedicated folders for pages, reusable UI components, styling, and utility functions.
- app/backend/: Contains the Node.js server code, which handles API logic and database interactions.
- tf/: Contains the code that defines the AWS infrastructure. This includes container hosting, firewalls, network routing and other services.
- docker-compose.yaml: Defines the services for the frontend, backend, and MongoDB database, ensuring the entire development environment can be managed with a single command.

To build a robust and secure home for our data, we constructed the MDAtlas platform as a ‘full-stack’ web application. The Atlas is designed to work together seamlessly and is neatly packaged using Docker technology, which ensures the entire system runs consistently and reliably.

The public-facing part of our platform the part users see and interact with is the frontend. We built this with React.js, which allows us to create a highly interactive and responsive experience. This is what powers the data tables, search bars, and visualisations, allowing for a smooth exploration of the data without constant page reloads. All the code for this user interface is neatly organised in its own app/frontend/ directory.

The backend was built with Node.js server in its own app/backend/directory. When a user filters for a specific gene or requests a dataset, the frontend sends this request to the backend. The backend then retrieves the necessary information from our database, performs any required calculations, and sends the results back to be displayed. It is the core logic that handles all the heavy lifting of data management and analysis.

The entire system (frontend, backend, and the MongoDB database) is managed by a single configuration file, “docker-compose.yaml”. This acts as the master blueprint, allowing us to run and manage all parts of the MDAtlas platform with a single, simple command.

When running in the cloud, the structure in the docker-compose.yaml is mirrored using AWS native services including ECS to host the application in a scalable and dynamic fashion. This is defined in Terraform scripts in the tf/ directory that are run by the CI/CD pipelines to set up and maintain the cloud infrastructure required to host the application.

#### Data Model Scheme

To ensure data within the Atlas is Findable, Accessible, Interoperable, and Reusable (FAIR) all information is organised according to a clear hierarchical data model. This ‘Data Model Scheme’ acts as our digital filing system, ensuring that every piece of information has a specific place and context. This structure connects every data file back to its biological context:

1. **Study (MDS):** The top-level project folder, representing an entire research project or initiative.
2. **Subject (MDSU):** Within each study are the de-identified participant information or subjects.
3. **Sample (MDSA):** The biological material collected. From each subject, a specific biological sample, like blood or tissue, is collected.
4. **Dataset (MDDA):** Finally, the raw data generated from a sample—such as the list of genetic variants, methylation levels, or gene expression counts—forms the dataset.

To keep track of all procedures, each entry is given a unique accession number (e.g., MDS00001), which acts like a permanent library catalogue number. This ensures that every piece of data, from the overarching study down to the individual gene, can be precisely located, referenced, and linked back to its source.

#### Interactive exploration and visualisation of multiomic data

The platform provides a rich interactive experience, leveraging a library of custom UI components including user friendly and responsive data tables, pagination controls, and dynamic charts for data visualisation. The state of the application is managed globally by Redux, ensuring a consistent and predictable user experience, with progress indicators and loading states providing real-time feedback during data-heavy operations. API interceptors are used to handle authentication seamlessly across the application. Users can export filtered data views as CSV files for local use.

#### Matching of rare variants across genomic datasets and generation of individual reports

Users can upload genomic data files, such as VCF and CSV formats, for comparison against MDAtlas database. The platform’s back-end analysis pipeline enables identification of variants within the uploaded data based on user-defined parameters (e.g AF).

#### Integration of MD genomic and epigenomic data

The primary function of the MDAtlas is multiomic integration. The platform’s analysis features allow for sophisticated filtering specific to each *omic*. Genomic data can be filtered according to the allelic frequency in gnomAD, population, gene name, specific position or genomic coordinate range, while the epigenomic data can be filtered according to gene name, specific position or range. This allows identification of rare (AF<0.05) and ultra-rare (AF<10^-3^) variants in the MD cohort per population, as well as methylation variant in the same genes, effectively integrating gene expression and protein pathogenicity data in MD individuals.

#### Understanding MD immune response and methylation profiles

Given that over 60% of MD cases exhibit an inflammatory phenotype, MDAtlas is specifically designed to facilitate research into its immunological basis of the disease. The RNASeq dataset provides raw counts from transcripts found in PBMC. In addition, the scRNAseq transcriptomic dataset offers detailed gene expression profiles for B cells, CD4^+^ and CD8^+^ T cells, NK cells and monocytes. This resource enables users to investigate how immune-related gene expression varies across different MD inflammatory phenotypes. Furthermore, methylation profiles have been generated from MD PBMCs, allowing users to retrieve DMC data alongside transcriptomic information to further analyse the immune landscape of MD.

#### Summary and future directions

MDAtlas provides a secure, collaborative platform for the global MD research community. successfully breaking down a major barrier to progress by integrating genomic, epigenomic, and transcriptomic datasets within a structured, searchable framework, it aims to accelerate the discovery of molecular markers and therapeutic targets.

Future development will focus on three key areas:

1. **Expanding Datasets:** Collaboration with global partners will allow for the enrichment of the database and will expand into new data types, such as proteomics, as they become available. The addition of **proteomic data** will provide the protein-level functions and metabolic state changes that result from the genetic and transcriptional variations we have catalogued.
2. **Advanced Analytics:** Predictive analyses with machine learning algorithms will be introduced. The goal is to develop integrative network models capable of identifying complex biomarker panels from the multiomic data. This will be paired with more sophisticated visualisation tools, such as interactive *cross-omic* dashboards where a user can see the relationship between a genetic variation, its methylation status, and its gene expression in one seamless view.
3. **Community Building:** Collaboration will be fostered by allowing users to securely share preliminary results and form research consortiums directly through the platform.

## CRediT author statement

K.B – Writing – original draft, Writing – Review and Editing, Data Curation, Visualisation, Software

P.C.G – Writing – original draft, Writing – Review and Editing, Data Curation, Visualisation

K.G – Writing – Review and Editing, Software

M.U – Review and Editing, Visualisation, Software

A.M.P.P – Review and Editing, Data Curation

S.M. – Review and Editing, Software

M.T.P. – Review and Editing, Data Curation

G.B.B. – Writing – Review and Editing, Data Curation

S.H.K – Review and Editing

R.A.F. – Review and Editing

D.B. – Writing – Review and Editing, Software

J.A.L.E – Conceptualisation, Funding Acquisition, Writing – original draft, Writing – Review and Editing, Supervision

## Human Ethics

The Human Ethics Research Committee (2023/HE000199) from The University of Sydney approved.

## Data Availability

The website can be found in the following URL: https://multiomic-md.sydney.edu.au/

## Acknowledgments

The Meniere Disease Multiomic Aggregation Consortium (MD-MAC) is a global crowdsourcing initiative to provide an open access resource with multiomic dataset about Meniere Disease (MD). List of participants in MD-MAC: Kiana Bagheri-Lotfabad, Pablo Cruz-Granados, Kurt Gobi, Muhammad Umair, Alberto M. Parra-Perez, Sayedali Mohseni, Anvitha Peddada, Maniratnam Meka, Ghulam Meeran, Maida Afzal, Zarif Raiyan Arefeen, Mai T. Pham, Giselle Bianco-Bortoletto, Sung Huhn Kim, Rick A. Friedman, Daniel Burn, Patricia Perez-Carpena, Jose A. Lopez-Escamez. We acknowledge all participant institutions that have provided de-identified data to build the datasets.

